# Automated optimization of deep brain stimulation parameters for modulating neuroimaging-based targets

**DOI:** 10.1101/2022.05.23.22275220

**Authors:** Mahsa Malekmohammadi, Richard Mustakos, Sameer Sheth, Nader Pouratian, Cameron C. McIntyre, Kelly R. Bijanki, Evangelia Tsolaki, Kevin Chiu, Meghan E. Robinson, Joshua A. Adkinson, Denise Oswalt, Stephen Carcieri

## Abstract

**Objective:** Therapeutic efficacy of deep brain stimulation (DBS) in both established and emerging indications, is highly dependent on accurate lead placement and optimized clinical programming. The latter relies on clinicians’ experience to search among available sets of stimulation parameters and can be limited by the time constraints of clinical practice. Recent innovations in device technology have expanded the number of possible electrode configurations and parameter sets available to clinicians, amplifying the challenge of time constraints. We hypothesize that patient specific neuroimaging data which can effectively assist the clinical programming using automated algorithms.

**Approach:** This paper introduces the DBS Illumina 3D algorithm as a tool which uses patient-specific imaging to find stimulation settings that optimizes activating a target area while minimizing the stimulation of areas outside the target that could result in unknown or undesired side effects. This approach utilizes preoperative neuroimaging data paired with the postoperative reconstruction of lead trajectory to search the available stimulation space and identify optimized stimulation parameters. We describe the application of this algorithm in three patients with treatment-resistant depression who underwent bilateral implantation of DBS in subcallosal cingulate cortex (SCC) and ventral capsule/ventral striatum (VC/VS) using tractography optimized targeting with an imaging defined target previously described.

**Main results:** Compared to the stimulation settings selected by the clinicians (informed by anatomy), stimulation settings produced by the algorithm achieved similar or greater target coverage, while producing a significantly smaller stimulation area that spills outside the target (P=0.002).

**Significance:** The DBS Illumina 3D algorithm is seamlessly integrated with the clinician programmer software and effectively and rapidly assists clinicians with the analysis of image based anatomy, and provides a starting point for the clinicians to search the highly complex stimulation parameter space and arrive at the stimulation settings that optimize activating a target area.

Clinical trial registration number: NCT 03437928

## Introduction

Deep brain stimulation (DBS) is an established and effective therapy for a diverse array of neuropathological conditions ranging from motor to cognitive and mood disorders. Standard clinical programming in DBS heavily relies on clinical experience and expertise and is often performed based on trial and error. During programming sessions, the clinician usually uses a programming device that communicates with the implanted pulse generator (IPG) to test many different stimulation settings, including electrode configuration, stimulation amplitude, pulse width, frequency and pulse patterns, and examine the patient for clinical response and the presence of potential side effects. This search aims to arrive at a stimulation setting that maximizes therapeutic benefit while minimizing the side effects [1,2]. Newest generations of DBS technology, such as those providing segmented leads and multiple independent current control (MICC), further expand the possible stimulation settings [3,4]. Although general guidelines for DBS programming are available [5–7], testing numerous stimulation settings is time consuming and exhaust clinicians, patients, and clinical resources.

Understanding the position of the electrode relative to the neuroanatomy could potentially facilitate both targeting and programming [8]. The value of imaging is highlighted by the significant increase in reports of equally efficacious outcomes with image-guided “asleep” implantation [9–11]. Patient-specific imaging data can also be paired with three-dimensional stimulation field models (SFMs) representing the volume of tissue activated (VTA) [12]. Pre-operative images are routinely used during DBS targeting and planning procedures. Anatomical segmentation of the pre-operative images, along with the reconstruction of the lead trajectory using post-operative imaging can provide unique information on the position of the lead with respect to the brain anatomy. Access to patient-specific anatomy, combined with information about stimulating various brain regions leading to clinical benefit or side effects, provides the ability to visualize the interaction of the stimulation models with respect to these structures and to potentially maximize the stimulation of beneficial areas and minimize the stimulation of regions that may adversely affect the patient. While SFM-guided programming is increasingly utilized in clinical practice, there is a need for tools which help the clinician optimize stimulation parameters to achieve maximal coverage of the targeted region by the estimated SFM.

In this paper we introduce and test a novel inverse programming algorithm, called the DBS Illumina 3D Algorithm which automates selection of optimized stimulation parameters for overlapping the SFM with a desired anatomical or functional target. The algorithm utilizes clinician-selected benefit and side effect regions from relevant anatomy to determine a set of suggested electrode configurations and stimulation amplitudes that maximize benefit-region coverage while minimizing the stimulation of regions of avoidance that could result in unknown or side effects. The user is able to prioritize both the importance of target region coverage versus total SFM volume and target region coverage versus side effect region stimulation. The DBS Illumina 3D Algorithm can be run multiple times to refine preferences and produce multiple sets of suggested initial stimulation parameters. These parameters can be used as a starting point to explore the options for treatment.

The target structure used in this report is from a set of clinician-chosen brain areas in a series of patients with treatment-resistant depression (TRD), using a target described in prior publications [13]. We compared clinician-chosen stimulation settings to algorithm-based settings and assessed the performance of algorithm. We demonstrate that the algorithm is fast and effective at placing the SFM to maximally overlap with a target structure, while minimizing the stimulation of non-target brain areas that could lead to unknown or undesired effects. Moreover, we demonstrate its performance in consistently stimulating the target across various pulse widths, emphasizing its utility in effectively searching the stimulation space based on the said criteria of maximally stimulating the target.

## Materials and Methods

### Patients

Individuals who participated in this study were all enrolled in the clinical trial (NCT 03437928) aimed at using a novel platform for therapy development based on elucidating the electrophysiological mechanisms underlying DBS for treatment resistant depression (TRD) [14,15]. These individuals were all diagnosed with treatment resistant major depressive disorder without psychotic features, and all provided written informed consent as approved by the Baylor College of Medicine IRB (H-43036) prior to participation. Details of the study participants are described elsewhere [14,15].

### Neuroimaging and implant procedure

Details of neuroimaging are provided previously [14,15]. T1-weighted anatomical imaging (MPRAGE; 1mm isotropic, TR/TE/TI=2400/2.24/1160; FOV=256; 208 slices; flip angle=8º) and Diffusion weighted imaging data (DWI) were acquired prior to surgical implantation of DBS. DWI data were acquired (1.5mm isotropic) with two phase encoding directions (anterior-to-posterior and posterior-to-anterior), 92 diffusion-sensitizing gradient directions, and 7 interleaved b=0 volumes. The diffusion-encoded volumes alternated between b=2000 and b=1000, with TR=3.2s, TE=87ms, TA=5:34 per scan, matrix 140×140×92, multi-slice acceleration=4 on a Siemens Prisma 3T scanner.

Study participants each underwent stereotactic implantation of four directional DBS leads (Cartesia™, Boston Scientific, Valencia, CA, USA), featuring 8 contacts in a 1-3-3-1 electrode configuration, where two leads were implanted in bilateral subcallosal cingulate cortex (SCC) and the other two were implanted in bilateral ventral striatum/ventral capsule (VC/VS). The target area for implantation, here referred to as the tractography-guided optimized target (TOT), was derived using pre-operative structural and diffusion weighted MR scans [13,16], Figure 1a. Each set of two leads were subcutaneously connected to a 16-contact Vercise Gevia rechargeable pulse generator (Boston Scientific, Valencia, CA, USA). The two pulse generators were implanted in the bilateral infraclavicular pockets of the chest wall.

**Figure 1-.**
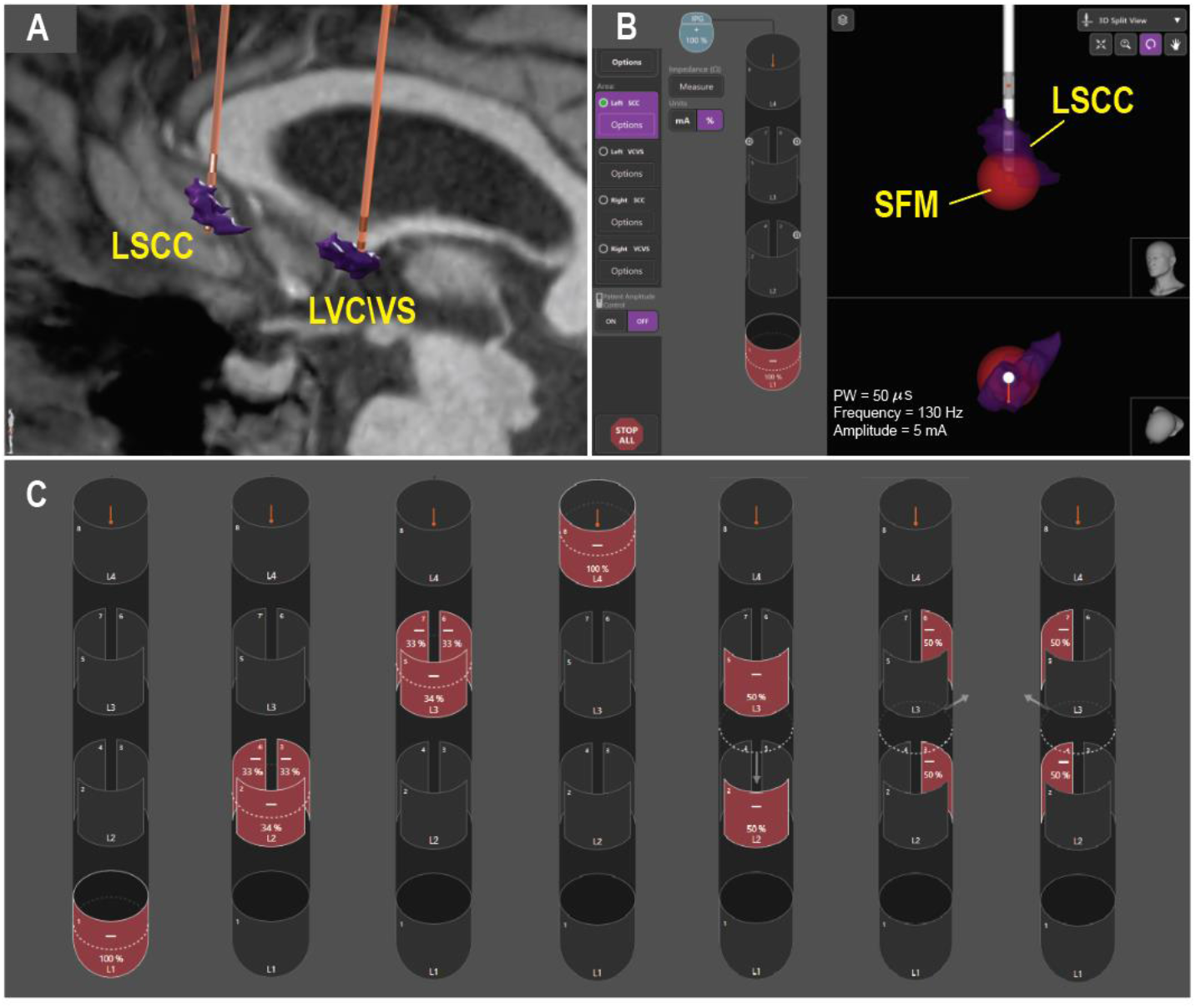
(A) Brain targets and reconstructed lead trajectories (B)Snapshot from Vercise Neural Navigator™ software showing an exemplary program and its corresponding SFM with respect to anatomy (target structure, TOT on LSCC (left subcallosal cingulate cortex) is shown in purple)(C) Stimulation configurations defined in the study protocol (i.e., Study sets), including four rings and three vertical stacks.

The post-implantation clinical CT scan was acquired on a Philips iCT 256 system, using a reconstruction diameter of 250mm, slice thickness of 0.67mm and a space between slices of 0.67mm, image size = 512×512, view size 1664×1236. Post-implantation CT scan was loaded in Brainlab Elements and co-registered to the preoperative structural scans using Image fusion module (Brainlab, Munich, Germany). The trajectory and orientation of each DBS lead was automatically extracted using the ‘Lead Localization’ module and was subsequently verified by visual inspection (Brainlab, Munich, Germany), Figure 1a [17]. Anatomical structures were defined with FSL (please see “Target area for Stimulation” below). The anatomical volumes and DBS lead trajectories were then transferred to Vercise Neural Navigator Software (Boston Scientific, Valencia, CA, USA) for clinical programming of the stimulation. The neural navigator enabled traditional manual adjustment to the DBS parameter settings, as well as experimental access to the Illumina algorithms for target-based optimization programming, Figure 1b (see next section for more details).

### Stimulation Field modeling

We use the term “fractionalization” to refer to the unique arrangement of current driven to each electrode, expressed as a percentage (fraction) of the total current. A fractionalization can include anodes (+), cathodes (−), and combinations of both polarities (Figure 1C, Table 1). To generate the stimulation field model (SFM) associated with each fractionalization, electric fields resulting from the stimulation setting are constructed as finite element models (FEM) using COMSOL Multiphysics software (COMSOL Inc., Burlington, MA, USA). The model consists of an insulating lead body having conducting electrodes, surrounded by an encapsulation layer, inside a cylinder of neural tissue. The neural tissue is modeled as isotropic and homogenous with conductivity of .2 S/m, and the encapsulation layer with a lower conductivity of .1 S/m. A multi-resolute mesh is created to encompass both the lead body and the encapsulation layer, with highest resolution at electrode-tissue interface and higher resolution in a region of interest (ROI) surrounding the electrode array versus the remaining volume. The scalar potentials at the mesh nodes are calculated and the model is solved once per electrode at unit current (1 mA).

**Table 1-.**
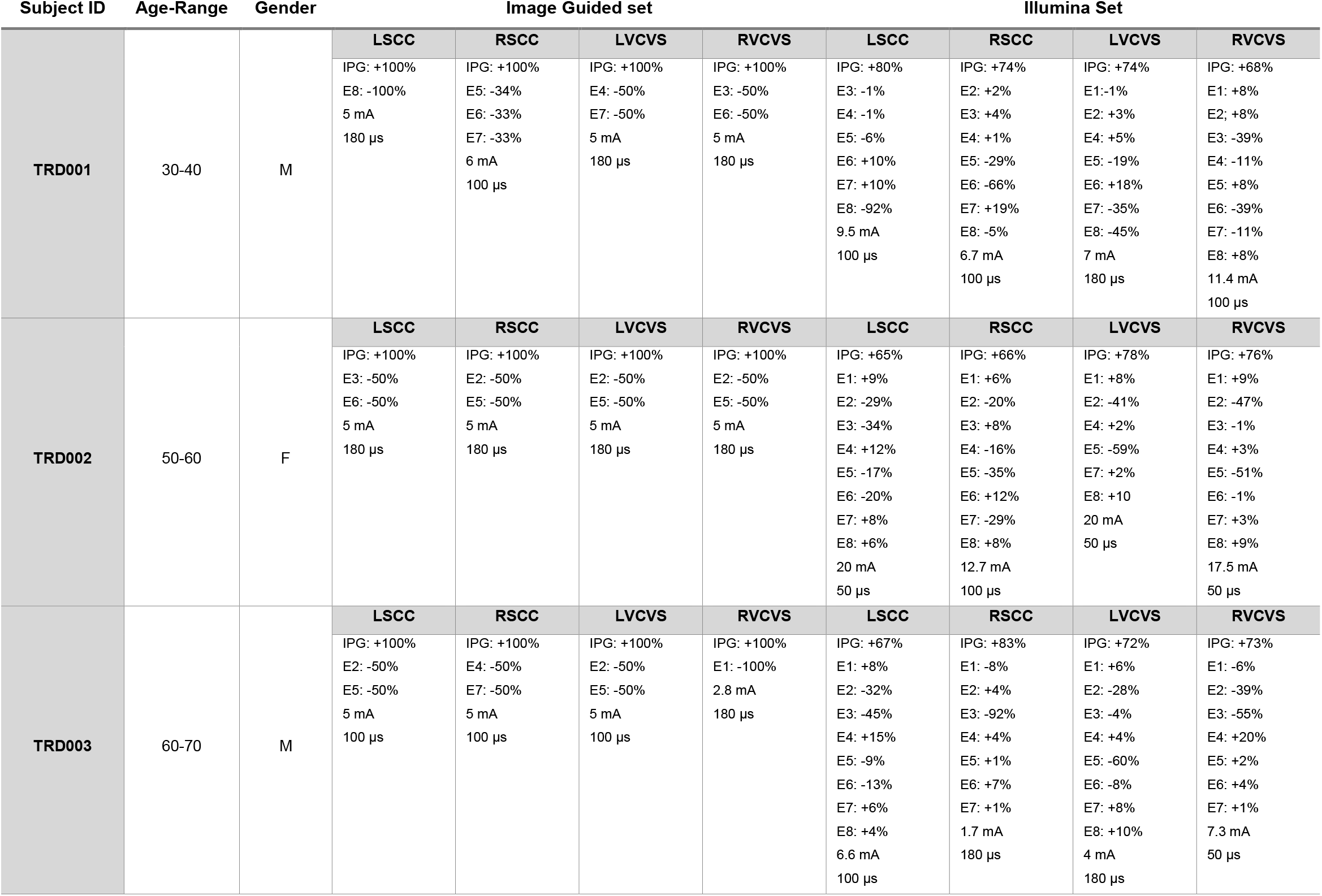
Demographics and Stimulation parameters for Image Guided and Illumina Sets. IPG stands for implanted pulse generator/Case. Curernt fractionalizations include anodes (+) and cathodes (−) along with % total current allocated to each contact (shown by letter “E” followed by the contact number)

The electric field results from the RoI are then exported from COMSOL and interpolated onto a regular grid of model axons that surround the DBS lead at 0.5 mm spacing (221-compartment, 21-node MRG myelinated axons of length of 10 mm and diameter of 5.7 μm [18]). The model consists of axons which are oriented orthogonally relative to the lead body and have identical behavior to a given stimulus. The response to each stimulus is computed by temporally scaling the potentials along the axon compartments using a waveform modeled on stimulator recordings to estimate the threshold current (‘Ith’, in mA) at which each axon in the grid fires an action potential from quiescence (NEURON, Yale, Version 7.3) [19]. A machine learning algorithm which takes features of the axon voltage profile as input and estimates axon’s response is trained on over 100 million axon simulations.

Basis files and the trained predictor are included in the Guide XT and Vercise Neural Navigator programmer software and can be integrated with the anatomical model of the patient. The output current amplitude thresholds for the axon models are iso-surfaced at the selected stimulation current amplitude. The resulting surface is displayed as the SFM. Exemplar SFM from Vercise Neual Navigator software is shown in Figure 1b.

### Target Area for Stimulation

Subject-specific TOT area within SCCwas defined in a semi-automated fashion, using methods previously described [13,16]. First, FSL probabilistic tractography [20] was performed to delineate the connectivity of SCC with patient-specific masks (ventral striatum (VS), uncinate fasciculus (UCF), anterior cingulate cortex (ACC) and bilateral medial prefrontal cortex (mFPC)) per hemisphere. A curvature threshold of 0.2 was used (approximately 80 degrees) for stopping streamline trajectories. The default 0.5mm voxel step length, 5000 samples and 2000 steps were used. Fibers with volume of fraction lower than 0.01 were discarded during tractography using the default value subsidiary fiber volume threshold. To avoid artifactual loops, streamlines that loop back on themselves were discarded. Using the -opd and -os2t flags 3D image files were created that contained the number of streamlines that reached each target voxel and seed segmentation maps to each target were derived where the value of each voxel corresponded to the number of streamlines seeded from that voxel reaching the target mask divided by the total number of streamlines (probability maps). Then the SCC probability maps for each target were smoothed using a Gaussian kernel (2 mm), multiplied on a voxel-by-voxel basis and then a high pass filter was applied (via thresholding) in order to include only voxels with probability higher than 10% of the maximum joint probability value.

Considering the activated tissue area surrounding the DBS electrode, the filtering of 2mm was considered the optimal option to simulate the area within SCC/VCVS that will present strong connectivity to each target in case of stimulation. Although the performed filtering approach might reduce the variability seen within the segmentation map between the voxels with low and high connectivity value, it highlights the areas within SCC/VCVS maps with higher probability of connectivity to each target. Finally, TOT within SCC was defined as the subregion with the highest joint probability of connectivity with all target areas. The same approach was followed to define the TOT within VCVS area using as targets the dorsolateral prefrontal cortex, nucleus accumbens, amygdala, medial and lateral orbitofrontal cortex.

### The DBS Illumina 3D Algorithm

The DBS Illumina 3D Algorithm (Boston Scientific, Valencia, CA USA) uses a metric optimization algorithm Bound Optimization by Quadratic Approximation (BOBQYA) [21]. The goal of the algorithm is to maximize stimulation of a target volume while staying within clinician-specified constraints. The algorithm incorporates the cost of increasing the size of the SFM, the cost of overlapping with avoidance volumes, including possible side effect regions, as well as stimulation safety limits.

The cost function, or metric, for the optimizer, for each fractionalization, is a weighted summation of the stimulated volumes for each structure (one target and one or more avoidance regions) and the SFM (background volume). The Target structure has a positive weight, and the avoidance structures and background have negative weights. For each fractionalization, the highest possible metric value is calculated, and the corresponding amplitude is determined. The clinician can specify one target region, zero or more avoidance region(s), the priority of not stimulating the avoidance regions (controlled by a slider to set ‘avoidance ratio’), and prioritization of reduced SFM volume (controlled by a slider to set the ‘background ratio’). The equation to calculate the optimized metric is therefore:

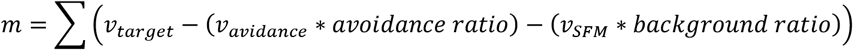

Where:

*m* = metric value

*v*_*target*_ = stimulated target volume in mm^3^

*v*_*avoidance*_ = stimulated avoidance volume in mm^3^

*v*_*SFM*_ = total SFM volume in mm^3^

In summary, the metric is the sum of the stimulated target volume (in mm^3^) minus the total volume of stimulated avoidance region (in mm^3^), weighted by the avoidance ratio, minus the total volume of background stimulation (in mm^3^), weighted by the background ratio. Where the avoidance ratio is the ratio of the cost (reduction in metric value) of stimulating avoidance region to the benefit (increase in metric value) of stimulating target region, and the background ratio is the ratio of the cost of stimulating background volume to the benefit of stimulating target region. The stimulated background volume is the same as the volume of the SFM.

The optimization algorithm is run once for each of two virtual electrode types (one equivalent to the ring electrodes on the lead, and one equivalent to the segmented electrodes on the lead).First, the optimizer is run using the rinig electrode, and a best solution is determined. If the lead is directional, the optimizer is run using the directional virtual electrode. As the optimization algorithm tests each virtual electrode’s position, the position is converted to a fractionalization on the real electrodes of the lead. For each fractionalization, the best metric among the possible amplitudes is compared to the metric of the current best solution. If the new metric is better than the previous best metric, the new metric, virtual electrode type, position, and derived amplitude are stored as the new best solution. When the optimization algorithm has met the stop conditions the best solution is returned and displayed for the clinician.

A virtual electrode shape defines a voltage field. For a gridded set of steering locations, the virtual electrode’s voltage field is transformed to that gridded location. Least squares fitting determines the fractionalization that produces the best fit between the real voltage field and the voltage field of the virtual electrode.

### Selection of Stimulation Parameters

The electrical stimulation was delivered as biphasic pulses of with passive recharge and at the frequency of 130 Hz. As part of the study protocol, SFMs from a series of pre-identified stimulation configurations were compared to find thosethat would maximally stimulate the target area. These pre-identified stimulation sets included four ring-mode configurations and 3 vertically stacked segments (Figure 1c), for each of the three pulse-width (PW) values of 50, 100 and 180 μs (total of 21 settings). Herein these sets are referred to as Study Sets. Per protocol, for each target, clinicians [NP ans SS] selected a final setting across all available Study Sets, based on the maximal volume of overlap with the target area. These final sets which we refer to as Image Guided Sets, represent anatomically-informed clinician-selected parameter sets that were subsequently used to program the patient as per study protocol (Table 1).

We subsequently used the DBS Illumina 3D Algorithm at each pulse width (50, 100 and 180 μs) to identify solutions for different selections of optimization cost, which we call here the Illumina Sets. In this application, the DBS Illumina 3D Algorithm only had one target area (i.e. TOT area corresponding to that lead) specified while no avoidance regions were selected.

### Statistical analysis

For each stimulation setting in the Study Sets, Image Guided Sets and Illumina Sets, we calculated the volume of resulting SFM by voxelizing the SFM isosurface and finding the number of voxels within the voxelized SFM, multiplied by the single voxel volume (1 mm^3^). The volume of overlap between the SFM and the target area, which is referred to as Fill Volume was similarly calculated by identifying the number of voxels that were common to both the SFM and the target area, multiplied by the single voxel volume (1 mm^3^). This volume was then normalized to the total target volume and converted to percentage to define % stimulated target volume. We also calculated the total volume of each SFM that spills outside the target area (i.e., Spill Volume) by subtracting the Fill Volume from the total volume of the SFM. These parameters were then shared with the clinicians (NP and SS) who compared various stimulation settings with respect to the objective criteria of activating the brain target while minimizing the stimulation of areas outside the target.

Statistically significant difference in Spill volume between Image guided Sets and Illumina Sets producing closest non-smaller target coverage was assessed using a non-parametric paired sample test (Wilcoxon signed rank test), P-values smaller than 0.5 were determined to be statistically significant. This specific comparison was motivated by the selection criteria for generation of Image Guided sets, to be maximally stimulating the target area.

Total charge deposited to the tissue per stimulation pulse was calculated by multiplying stimulation amplitude and pulse-width. Total charge was used to establish decision guiding criteria for selection of stimulation parameters across various combinations.

## Results

Stimulation settings were evaluated in four targets in each of the three individuals enrolled in this study (Table 1). After performing image processing steps described in the methods sections, brain targets (i.e., TOT areas) and lead trajectories were defined for each lead and Study sets and Illumina sets were generated according to the study protocol.

We first generated SFMs from Illumina sets, Study sets, and Image Guided sets to identify which group provided optimized target coverage (i.e. Fill) versus spill volume. Example in Figure 2a shows a sample from Study sets from Left SCC in TRD003 (top panel, highlighted by a red filled square in Figure 2b) and a comparable sample from Illumina sets (bottom panel, highlighted by filled circle in Figure 2b). Image Guided sets were selected from Study sets as those providing maximal volume of target coverage.

**Figure 2-.**
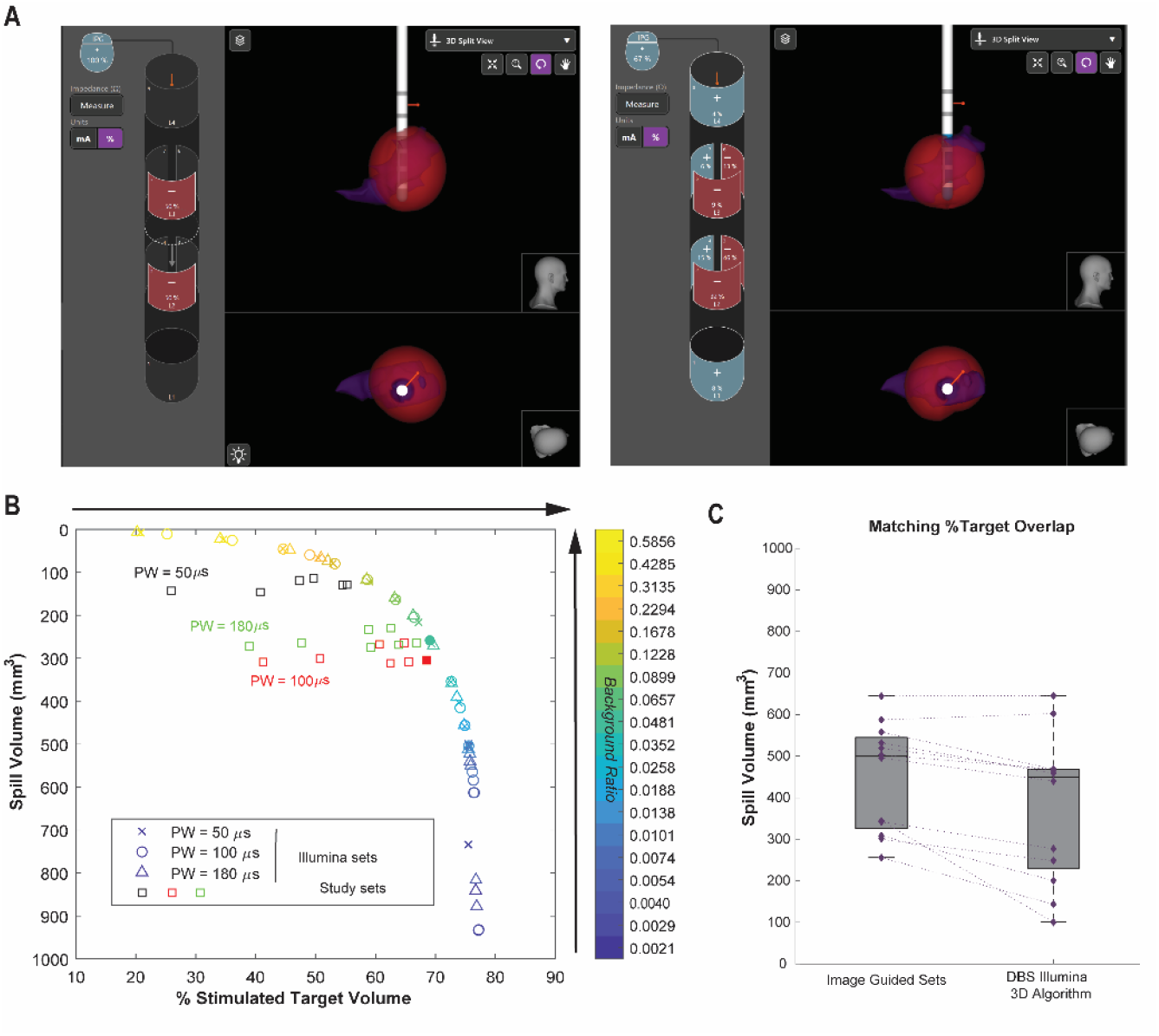
Study sets and Illumina sets. (A) Left panel shows exemplary stimulation setting from one of the Study sets at PW = 100 μS that was selected as the Image Guided set (highlighted in by filled red square in panel B) and how a matching output form Illumina sets (right panel) produces more complex current fractionalization (including multipolar stimulation) to optimize filling the target area (i.e., for similar or greater target coverage, produces less Spill Volume, see panel (B)). SFM is shown in red and brain target area shown in purple (left SCC). (B) a series of SFMs generated from Illumina (different symbols: ×, º and Δ for PW = 50, 100 and 180 μS respectively) and corresponding series of SFMs generated from Study sets (symbol: □, different colors used for PW = 50, 100 and 180 μS) are compared based on Spill volume vs % Stimulated target Volume. As shown by arrows outside the graph, larger % Stimulated Target Volume and Smaller Spill volume are desired. (C) Boxplots comparing Spill Volume between Image Guided sets and their paired Illumina sets, identified by matching the % stimulated target volume, such that Illumina Sets were providing closest non-smaller target coverage to the Image Guided sets. Paired sample Wilcoxon signed rank test indicated a significant difference between the two group (P = 0.002). Illumina results provided significantly smaller Spill Volume.

DBS Illumina 3D Algorithm allowed the user to move a slider to adjust the Background Ratio which establishes the cost of stimulating the target area versus stimulating the area outside the target (Color bar in Figure 2b). Hence, a series of Illumina sets were generated for each pulsewidth. For each target, one single Illumina Set, that provided closest non-smaller Fill Volume to its corresponding Image Guided set was selected to be compared against Image Guided sets (Table 1). We then performed a series of comparisons between these groups of settings and present our findings below

### The DBS Illumina 3D algorithm found stimulation settings with optimized target coverage

Detailed comparison across Illumina sets for different values of Background Ratio and all the study sets, indicated that for a given Spill Volume, across three pulse-widths, the algorithm identifies solutions with Fill Volumes at least on par but usually greater that the largest fill volumes seen with the Study set (Figure 2b). Conversely, for similar values of target coverage (% stimulated target volume), SFMs generated from Illumina sets, provide at least the same or more often smaller Spill Volume compared to Study sets. These findings were consistently observed across all targets in all study participants. Statistical comparison between Image Guided Set, which represent what clinicians selected among Study sets to maximize target coverage, and their matching Illumina Set (i.e., those providing closest non-smaller Fill Volume), indicated that Illumina Sets provided significantly smaller Spill Volume (Wilcoxon signed rank test, P= 0.002, Figure 2C).

### Illumina sets showed similar target coverage vs spill volume across various pulse widths

Flexibility of DBS Illumina 3D algorithm allows for identifying various stimulation sets per pulsewidth via selecting different values for Background Ratio (Color bar in Figure 2b). In all of the 12 targets, for each Background ratio, the stimulation sets offered similar target coverage across three PWs of 50, 100 and 180 μs (Figure 2b). To create a better understanding of this behavior, we expanded the selection of PW to include values between 20 and 180 μs (at 10 μs steps) and found corresponding Illumina results. We then calculated % Stimulated target Volume for all the resulting stimulation sets (Figure 3a). When collapsed across the PW dimension, we confirmed that for every selected Background Ratio, variability in % Stimulated target Volume is small (standard deviation: 0.28-3.1 % Stimulated Target Volume) across all PWs (between 20 and 180 μs), Figure 2b.

**Figure 3-.**
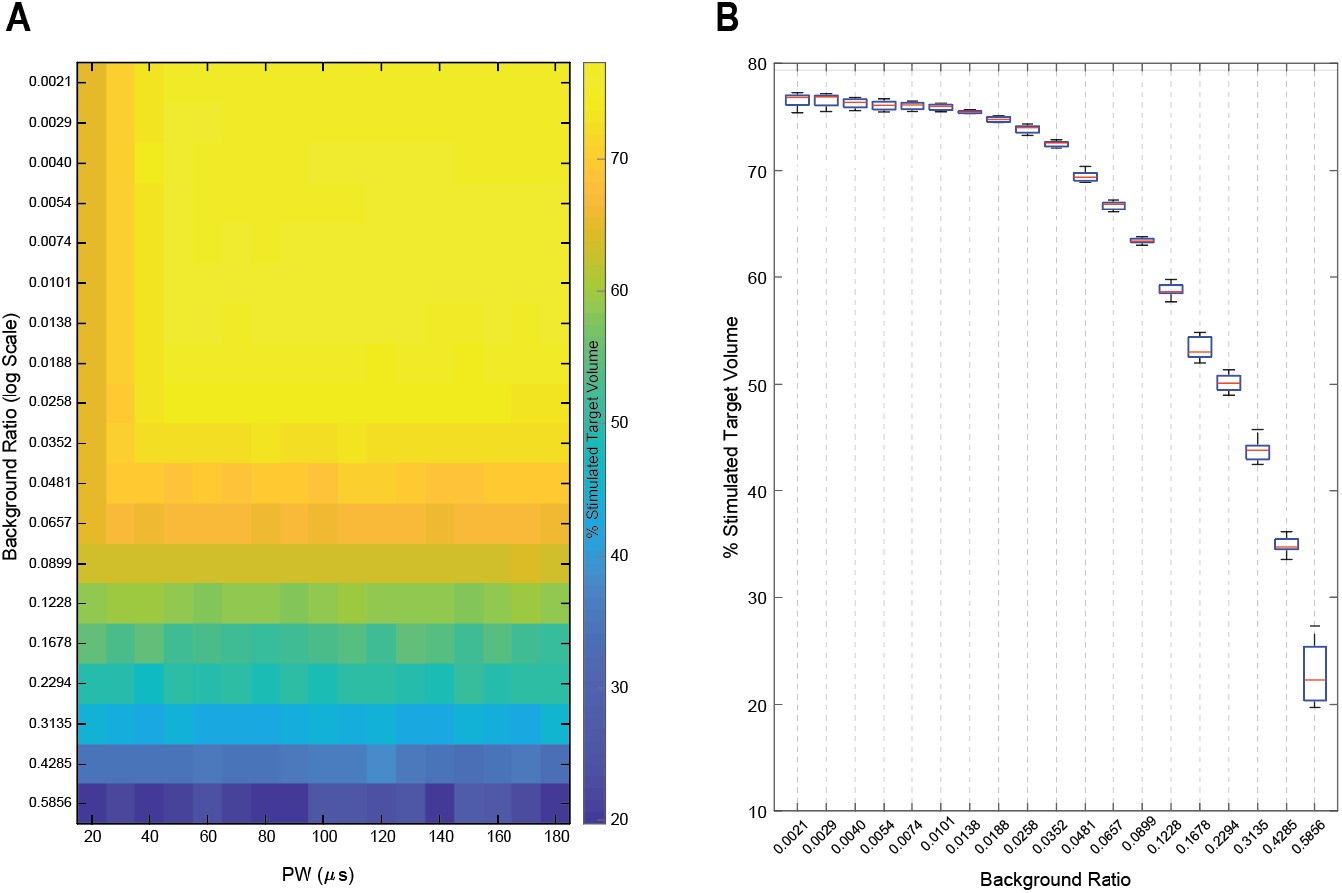
DBS Illumina 3D algorithm shows consistent behavior across various selections of PW and background Ratios (data presented from the same target as presented in Figure 2. TRD003 and LSCC (A) Stimulated target volume for Illumina results generated for various PW and background ratios. (B) Once the data is collapsed across the PW dimension, Illumina produces near similar % stimulated target volume for different selections of Background ratio.

Given the consistent behavior of the DBS Illumina 3D algorithm in providing similar target coverage vs Spill profile across various pulse widths, it is important to provide potential guidance to the user clinicians to assist with selection of appropriate background ratio. This selection can potentially be informed by gaining greater quantitative understanding of the adverse effects related to stimulation of avoidance regions. Other factors such as efficiency of stimulation can and should also be considered. For example, holding constant the amount of total energy or charge delivered per pulse, which affects both the rate of battery usage and the total volume of the produced SFM, can be used as a technique for making comparisons across different combinations of DBS parameters [22]. To provide an example, we have performed some basic analysis to create SFM families that provide a constant charge deposited to the tissue for various selections of PW and amplitude (Figure 4).

**Figure 4-.**
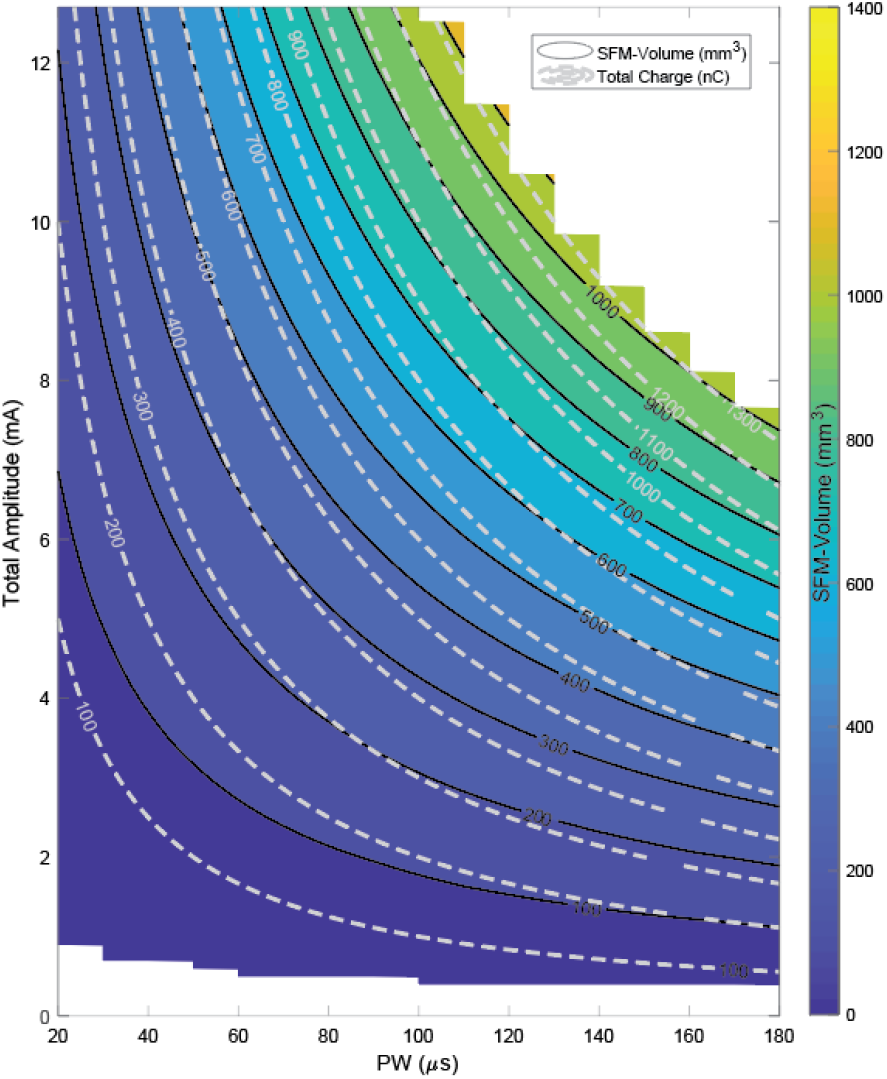
Example SFM volume for a fixed fractionalization (E1: 100%) as a function of stimulation amplitude (mA) and pulse width (PW, μs). Solid Black and dashed gray contours (i.e. isolines) indicate constant volume and constant total charge produced by these SFMs.

## Discussion

DBS has become an established therapy for the management of movement disorders (such as Parkinson disease, essential tremor and dystonia) and is being actively investigated for emerging new indications such as treatment resistant depression, and other neuropsychiatric indications. Effective symptom control for both established and emerging applications is directly related to identifying the right brain area to implant the lead and subsequently to program the lead to optimally stimulate the designated target area. Once DBS lead implantation has been finalized, programming adjustment is the only factor that could affect the therapy, highlighting the crucial role of programming, especially when the DBS electrodes are sub optimally placed at the border of the intended target structure.

Over the past few years, there has been an emerging interest to develop tools that use neuroimaging data to assist with identification of stimulation settings that could assist with DBS programming [23,24]. Nevertheless, these advances have been mostly restricted to highly specialized centers with a strong computational background and are not approved for clinical use. Efforts are being undertaken to develop user-friendly and clinically validated software which may foster a more refined search strategy for identifying stimulation settings that would optimize activating a pre-specified target area.

Here we have introduced and characterized the DBS Illumina 3D Algorithm as a novel inverse programming algorithm which automates the search with providing solutions in a matter of seconds, and generally out-performs human experts at matching stimulation settings to patient-specific anatomical targets based on imaging data. Our findings suggest that the DBS Illumina 3D Algorithm can be effectively utilized to assist with identifying stimulation parameters that optimizes activating a target volume with controlled spill outside the target. The algorithm provided consistent performance across the wide range of stimulation pulse widths including those used in routine clinical practice while maintaining flexibility based on the user’s clinical judgment and needs, by weighting stimulation of a target area versus stimulating outside the target area (background ratio) to rapidly provide a series of initial stimulation settings that can be subsequently evaluated by clinical tests. Regarding the algorithm design, the idea of trading off between stimulating the target and stimulating avoidance volumes is based on the observation that fully stimulating the target may often by impossible without some activation of other structures, including avoidance volumes. Therefore, a trade-off is required when deciding whether to increase the volume of tissue activation—an increase in VTA will often increase both the activation of target volume and the activation of undesired areas, leaving the clinician with the need to control the trade-off. It is possible that volumes that are described may require a threshold of overlap with the stimulation field, and that the decision of how much overlap is allowed should be left to the clinician’s judgement.

Our philospphy in designing the objective function used in the DBS Illumina 3D algorithm is informed by previous work [25,26] roughly the same as that used in Pena et al, 2017, with SFM volume being a surrogate for power, and the optimizer in Illumina DBS maximizing the metric, as opposed to minimizing it [27]. More plainly, Pena et al allow the stimulation of avoidance region. The ratio of acceptable avoidance volume stimulation to target volume stimulation is hard coded to 2.0, meaning stimulating an additional 2 units in the target allows the stimulation of one unit in the avoidance region, and the user has no method of specifying some other ratio. Our philosophy behind allowing the user to select how sensitive they want the algorithm to be in regard to increasing SFM size and Avoidance region overlap is one of pragmatism. While specific values could have been chosen for the algorithm, our belief is that the clinician will have preferences and gain experience allowing them to determine what they think is best, and that this is better than preselecting a set of hard coded ratios.

Although further work is needed to determine whether this technique results in providing superior therapeutic benefits for DBS patients, we believe the DBS Illumina 3D Algorithm holds promise to facilitate image-guided selection of stimulation parameters and significantly reduce the time and energy required for trial and error based clinical testing. Our current work focused on application of the DBS Illumina 3D Algorithm in newly proposed targets that are being actively investigated for DBS in treatment resistant depression, as an example. However, the technique is agnostic to the target, as long as it can be defined radiographically relative to DBS lead and contact positions. We purposefully avoid defining a subthalamic or pallidal target of stimulation, as that in and of itself remains controversial. More specifically, different research groups have different ways to define the most clinically appropriate STN/GPi based target areas. Therefore, our intent was to focus on the optimization of stimulation parameters based on any target of interest.

## Limitations

Our work focuses on application of the DBS Illumina 3D Algorithm in newly proposed targets that are being actively investigated for DBS in treatment resistant depression, as an example. Given the nature of this study to interrogate tractography optimized targates (TOTs) in TRD, the focus of study was to maximally activate the target area while avoiding the stimulation of areas outside the target. Fture work is needed to explore application of DBS Illumina 3D algorithm in more complex scenarios incorporating specific avoidance areas.

, The technique used in this study via DBS Illumina 3D algorithm is agnostic to the target, as long as it can be defined radiographically relative to DBS lead and contact positions. We purposefully avoid defining a subthalamic or pallidal target of stimulation, as that in and of itself remains controversial. More specifically, different research groups have different ways to define the most clinically appropriate STN/GPi based target areas. Therefore, our intent was to focus on the optimization of stimulation parameters based on any target of interest.

We further acknlowdge that the usage of homogenous and isotropic neural tissue as well as selection of a single fiber diameter neuron in our SFM modeling may limit the DBS Illumina 3D Algorthm in providing final and anatomy specific stimulation solutions. Although antomy-specifc modling will be more sensitive and certainly improve the accuracy of personalized treatment optimization; Recognizing these limitations, our goal in creating the DBS Illumina 3D algorithm was to facilitate searching the large and complex treatment space, which becomes increasingly important as newer generations of stimulation systems become available (systems capable of controlling the fractionalization at very fine resolutions and with mixed polarity, as well as DBS leads with complex segmented and directional designs). The DBS Illumina 3D algorithm aims to provide anatomically-informed “starting” point. This optimized starting solution needs to be further refined, most likely accompanied by direct clinical evalution of the patient.

## Conclusion

Algorithm-guided clinical programming of DBS that uses relative position of the electrodes with respect to anatomical or functional neuroimaging targets may be an effective approach to replace traditional monopolar review. Seamless integration of this algorithm with the DBS surgical workflow (from targeting to programming) can enable larger group of clinicians with variable experience and expertise in neuroimaging to rapidly Interrogate the large space of possible stimulation settings and reduce time needed for programming. Although establishing therapeutic efficacy of this set of brain targets and their application in TRD has not been the focus of this work, we argue that this application could potentially lay the ground work for other novel indications to facilitate testing of imaging-informed selection of stimulation parameters and enable interrogation of different targeting strategies, including those informed by aggregation of prior information derived from group SFM maps, or functional or structural connectivity patterns.

## Data Availability

All data produced in the present study are available upon reasonable request to the authors

## Acknowledgements

Research reported in this publication was supported by the National Institute of Neurological Diseases and Stroke of the National Institutes of Health under Award Number UH3 NS103549. The content is solely the responsibility of the authors does not necessarily represent the official views of the National Institutes of Health.

MM, RM and SC are employees of Boston Scientific Corporation. KC is an employee of Brainlab corporation. CCM is a paid consultant for Boston Scientific Neuromodulation, receives royalties from Hologram Consultants, Neuros Medical, Qr8 Health, and is a shareholder in the following companies: Hologram Consultants, Surgical Information Sciences, CereGate, Autonomic Technologies, Cardionomic, Enspire DBS. NP is a consultant for Abbott, Boston Scientific, Sensoria Therapeutic, and Second Sight Medical Products. SAS is a consultant for Boston Scientific, Zimmer Biomet, Neuropace, Abbott, and Koh Young.

## References

[1] Anon Deep Brain Stimulation Programming: Mechanisms, Principles, and Practice - Erwin B. Montgomery, Jr. - Google Books

[2] Oliveira Godeiro C de, Moro E and Montgomery E B 2020 Programming: General Aspects Fundam. Clin. Deep Brain Stimul. 93–125

[3] Dembek T A, Reker P, Visser-Vandewalle V, Wirths J, Treuer H, Klehr M, Roediger J, Dafsari H S, Barbe M T and Timmermann L 2017 Directional DBS increases side-effect thresholds—A prospective, double-blind trial Mov. Disord. 32 1380–8

[4] Kramme J, Dembek T A, Treuer H, Dafsari H S, Barbe M T, Wirths J and Visser-Vandewalle V 2021 Potentials and Limitations of Directional Deep Brain Stimulation: A Simulation Approach Stereotact. Funct. Neurosurg. 99 65–74

[5] J V, E M and R P 2006 Basic algorithms for the programming of deep brain stimulation in Parkinson’s disease Mov. Disord. 21 Suppl 14

[6] M P, AM L, N K, R P M and A F 2016 Programming Deep Brain Stimulation for Parkinson’s Disease: The Toronto Western Hospital Algorithms Brain Stimul. 9 425–37

[7] Koeglsperger T, Palleis C, Hell F, Mehrkens J H and Bötzel K 2019 Deep Brain Stimulation Programming for Movement Disorders: Current Concepts and Evidence-Based Strategies Front. Neurol. 10 410

[8] Waldthaler J, Bopp M, Kühn N, Bacara B, Keuler M, Gjorgjevski M, Carl B, Timmermann L, Nimsky C and Pedrosa D J 2021 Imaging-based programming of subthalamic nucleus deep brain stimulation in Parkinson’s disease Brain Stimul. 14 1109–17

[9] Brodsky M A, Anderson S, Murchison C, Seier M, Wilhelm J, Vederman A and Burchiel K J 2017 Clinical outcomes of asleep vs awake deep brain stimulation for Parkinson disease Neurology 89 1944–50

[10] Jin H, Gong S, Tao Y, Huo H, Sun X, Song D, Xu M, Xu Z, Liu Y, Wang S, Yuan L, Wang T, Song W and Pan H 2020 A comparative study of asleep and awake deep brain stimulation robot-assisted surgery for Parkinson’s disease npj Park. Dis. 2020 61 6 1–7

[11] Engelhardt J, Caire F, Damon-Perrière N, Guehl D, Branchard O, Auzou N, Tison F, Meissner W G, Krim E, Bannier S, Bénard A, Sitta R, Fontaine D, Hoarau X, Burbaud P and Cuny E 2021 A Phase 2 Randomized Trial of Asleep versus Awake Subthalamic Nucleus Deep Brain Stimulation for Parkinson’s Disease Stereotact. Funct. Neurosurg. 99 230–40

[12] Vedam-Mai V, Deisseroth K, Giordano J, Lazaro-Munoz G, Chiong W, Suthana N, Langevin J-P, Gill J, Goodman W, Provenza N R, Halpern C H, Shivacharan R S, Cunningham T N, Sheth S A, Pouratian N, Scangos K W, Mayberg H S, Horn A, Johnson K A, Butson C R, Gilron R, de Hemptinne C, Wilt R, Yaroshinsky M, Little S, Starr P, Worrell G, Shirvalkar P, Chang E, Volkmann J, Muthuraman M, Groppa S, Kühn A A, Li L, Johnson M, Otto K J, Raike R, Goetz S, Wu C, Silburn P, Cheeran B, Pathak Y J, Malekmohammadi M, Gunduz A, Wong J K, Cernera S, Hu W, Wagle Shukla A, Ramirez-Zamora A, Deeb W, Patterson A, Foote K D and Okun M S 2021 Proceedings of the Eighth Annual Deep Brain Stimulation Think Tank: Advances in Optogenetics, Ethical Issues Affecting DBS Research, Neuromodulatory Approaches for Depression, Adaptive Neurostimulation, and Emerging DBS Technologies Front. Hum. Neurosci. 0 169

[13] Tsolaki E, Espinoza R and Pouratian N 2017 Using probabilistic tractography to target the subcallosal cingulate cortex in patients with treatment resistant depression Psychiatry Res. 261 72

[14] Allawala A, Bijanki K R, Goodman W, Cohn J F, Viswanathan A, Yoshor D, Borton D A, Pouratian N and Sheth S A 2021 A Novel Framework for Network-Targeted Neuropsychiatric Deep Brain Stimulation Neurosurgery 89 E116–21

[15] Sheth S A, Bijanki K R, Metzger B, Allawala A, Pirtle V, Adkinson J A, Myers J, Mathura R K, Oswalt D, Tsolaki E, Xiao J, Noecker A, Strutt A M, Cohn J F, McIntyre C C, Mathew S J, Borton D, Goodman W and Pouratian N 2021 Deep brain stimulation for depression informed by intracranial recordings Biol. Psychiatry 0

[16] Tsolaki E, Sheth S A and Pouratian N 2021 Variability of white matter anatomy in the subcallosal cingulate area Hum. Brain Mapp. 42 2005

[17] Hellerbach A, Dembek T A, Hoevels M, Holz J A, Gierich A, Luyken K, Barbe M T, Wirths J, Visser-Vandewalle V and Treuer H 2018 DiODe: Directional orientation detection of segmented deep brain stimulation leads: A sequential algorithm based on CT imaging Stereotact. Funct. Neurosurg. 96 335–41

[18] McIntyre C C, Richardson A G and Grill W M 2002 Modeling the Excitability of Mammalian Nerve Fibers: Influence of Afterpotentials on the Recovery Cycle https://doi.org/10.1152/jn.00353.2001 87 995–1006

[19] Hines M L and Carnevale N T 1997 The NEURON Simulation Environment Neural Comput. 9 1179–209

[20] Behrens T E J, Berg H J, Jbabdi S, Rushworth M F S and Woolrich M W 2007 Probabilistic diffusion tractography with multiple fibre orientations: What can we gain? Neuroimage 34 144–55

[21] Powell M 2009 The BOBYQA algorithm for bound constrained optimization without derivatives NA Rep. NA2009/06 39

[22] Moro E, Esselink R J A, Xie J, Hommel M, Benabid A L and Pollak P 2002 The impact on Parkinson’s disease of electrical parameter settings in STN stimulation Neurology 59 706–13

[23] Horn A and Kühn A A 2015 Lead-DBS: A toolbox for deep brain stimulation electrode localizations and visualizations Neuroimage 107 127–35

[24] Horn A, Li N, Dembek T A, Kappel A, Boulay C, Ewert S, Tietze A, Husch A, Perera T, Neumann W-J, Reisert M, Si H, Oostenveld R, Rorden C, Yeh F-C, Fang Q, Herrington T M, Vorwerk J and Kühn A A 2019 Lead-DBS v2: Towards a comprehensive pipeline for deep brain stimulation imaging Neuroimage 184 293–316

[25] Anderson D N, Osting B, Vorwerk J, Dorval A D and Butson C R 2018 Optimized programming algorithm for cylindrical and directional deep brain stimulation electrodes J. Neural Eng. 15

[26] Vorwerk J, Brock A A, Anderson D N, Rolston J D and Butson C R 2019 A retrospective evaluation of automated optimization of deep brain stimulation parameters J. Neural Eng. 16

[27] Peña E, Zhang S, Deyo S, Xiao Y and Johnson M D 2017 Particle Swarm Optimization for Programming Deep Brain Stimulation Arrays J. Neural Eng. 14 016014

